# Diabetes Self-Management Education and Support Models and Their Impact on Clinical and Psychosocial Outcomes: A Systematic Review

**DOI:** 10.1101/2025.09.13.25335687

**Authors:** Vina Putri Patandung, Moses Glorino Rumambo Pandin

## Abstract

**Background:** Diabetes self-management education and support (DSMES) is central to glycemic control and complication prevention. The aim of this review is to synthesize evidence from six selected studies on various DSMES models (collaborative/cultural, Kolb’s learning style– based, telemedicine, community or group-based interventions, and foot ulcer prevention training) and to situate these findings within the current literature (2020–2025).

**Methods:** The literature screening and reporting process followed PRISMA guidelines. Six articles analyzed were obtained from user-uploaded files (randomized controlled trials, experimental, and quasi-experimental studies). Data were systematically extracted, including methods, population, duration, and main outcomes. The discussion integrates evidence from 20 Scopus-indexed articles (2020–2025).

**Results:** All six studies demonstrated significant improvements in at least one outcome (self-care behavior, HbA1c, distress, or foot complication prevention) following DSMES interventions, although long-term effects varied. Culturally tailored programs, sustained support through telemedicine, and learning style–adapted approaches reported promising clinical and psychosocial outcomes.

**Conclusion:** Structured DSMES, particularly when personalized, culturally adapted, and supported by telehealth or continuous follow-up, is effective in improving self-care and several short-term clinical outcomes. Recommendations include standardization of core DSMES components, emphasis on regular follow-up, and long-term studies with larger sample sizes.

## INTRODUCTION

Type 2 diabetes mellitus (T2DM) is a major global health problem with rising prevalence driven by demographic transitions, urbanization, and lifestyle changes. The International Diabetes Federation (IDF) estimated that 537 million people were living with diabetes in 2021, a number projected to reach 783 million by 2045, with T2DM accounting for the majority of cases. The condition is associated with long-term complications, including cardiovascular disease, nephropathy, neuropathy, and retinopathy, which reduce quality of life and increase healthcare costs (Sun *et al*., 2022).

Diabetes Self-Management Education and Support (DSMES) has become a cornerstone of T2DM care. DSMES is defined as a collaborative process that helps patients acquire knowledge, skills, and motivation to effectively manage their condition. Various models have been developed, including group-based sessions, individualized interventions, culturally adapted programs, and digital or telemedicine approaches (Ernawati, Suharto and Dewi, 2015; Siminerio *et al*., 2023).

Evidence suggests that DSMES improves glycemic control, enhances self-care behaviors, and reduces diabetes distress. For instance, Bagchegi *et al* (2021) found that education grounded in Kolb’s experiential learning theory improved self-care among elderly patients with T2DM, while Anjali *et al* (2023) demonstrated the effectiveness of brief face-to-face education combined with instant messaging follow-up in reducing HbA1c levels. Cultural adaptation also plays an important role in the success of DSMES. Hempler *et al* (2023) showed that programs co-developed with immigrant communities by integrating language, values, and cultural practices were effective in improving both HbA1c and self-management behaviors. Such findings highlight the importance of context-sensitive interventions in diabetes education (Zhang *et al*., 2025).

Alongside personalization and cultural sensitivity, technology-based delivery has expanded access to DSMES. Tele-DSMES, as reported by Siminerio *et al* (2023) through the TREAT-ON model, achieved outcomes comparable to traditional care for rural populations, with high patient satisfaction. Recent meta-analyses further confirm the effectiveness of telemedicine-based DSMES, particularly during the COVID-19 pandemic (Chiaranai *et al*., 2024).

Despite promising outcomes, heterogeneity in intervention design, duration, and intensity remains a challenge, with some effects diminishing over time without sustained support (Liu *et al*., 2024). Therefore, this systematic review aims to evaluate the effectiveness of different DSMES models reported in recent studies (2018–2023), compare them with contemporary literature (2020–2025), and identify key factors for optimizing DSMES in clinical practice.

## METHODS

### Study Design

The process of literature screening, inclusion, and reporting followed the PRISMA (Preferred Reporting Items for Systematic Reviews and Meta-Analyses) guidelines. The search was conducted in Scopus, PubMed, Web of Science (WoS), Science Direct, and EBSCO databases from 2020 to 2025.

**Figure 1.**
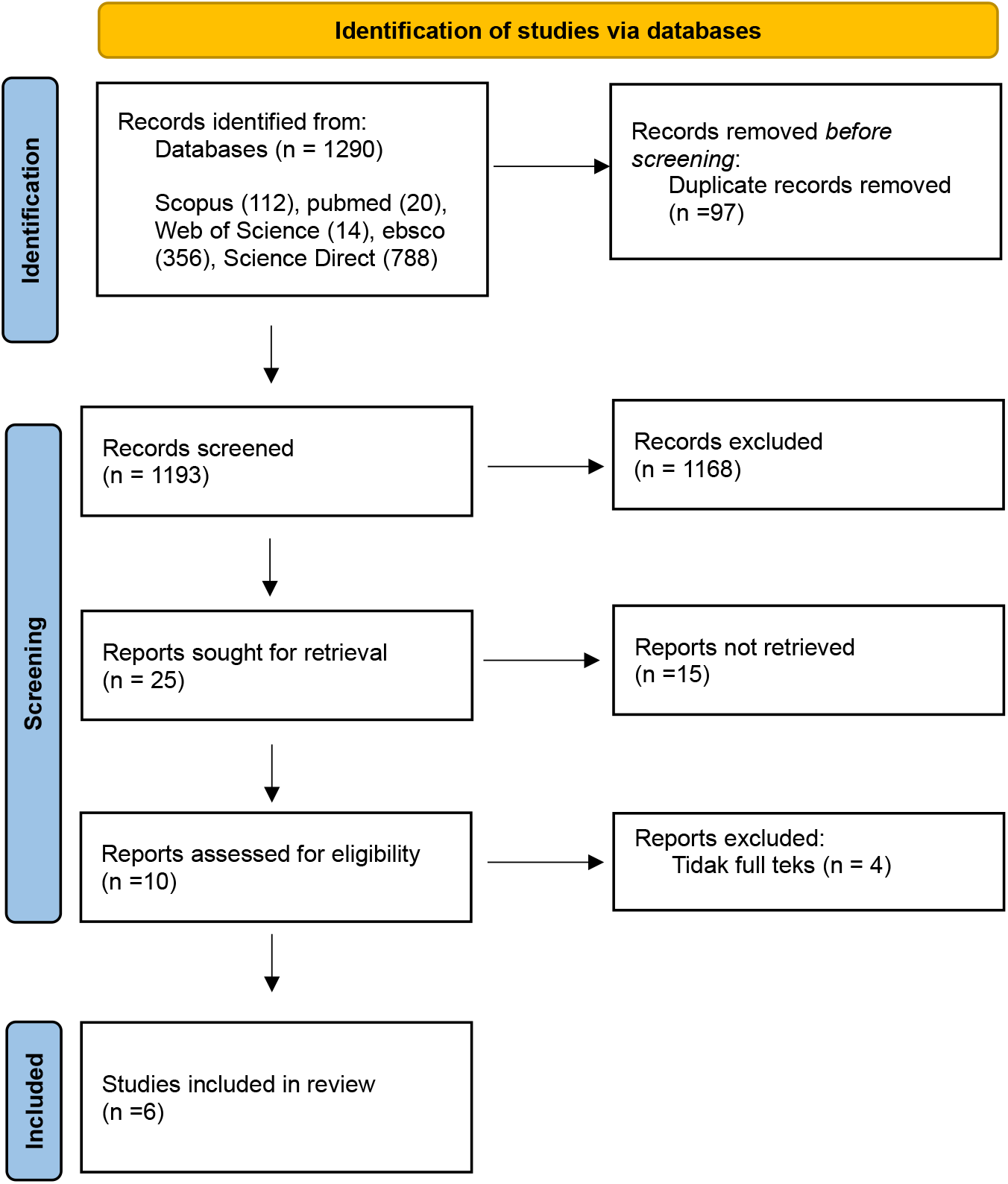
The identification, screening, and inclusion procedures of the studies were available in Scopus, PubMed, Web of Science (WoS), Science Direct, and EBSCO databases.

### Inclusion and Exclusion Criteria

The inclusion criteria for this review were defined to capture studies that specifically evaluated the effectiveness of Diabetes Self-Management Education and Support (DSMES) interventions. Eligible studies were those that implemented DSMES in various formats, including face-to-face sessions, group-based programs, telehealth or technology-supported approaches, culturally tailored interventions, or education designed around specific learning models. To ensure relevance, only studies that reported clinical, psychosocial, or self-care outcomes among adults or older adults with type 2 diabetes were considered. Regarding study design, randomized controlled trials (RCTs), quasi-experimental studies, pre–post evaluations, and implementation research were included. Furthermore, the review focused on publications between 2018 and 2025, with particular emphasis on the 2020–2025 period to provide an up-to-date discussion. Articles published in either English or Indonesian were eligible. Importantly, six articles obtained from the uploaded files served as the primary data sources and were synthesized in the Results section.

Exclusion criteria were applied to ensure the rigor and focus of the review. Studies were excluded if they were non-interventional in nature, such as descriptive reports, editorials, or commentaries, that did not evaluate outcomes. Similarly, studies focusing on type 1 diabetes in children were not considered, unless the intervention specifically targeted adult caregivers in the context of DSMES. Finally, duplicate publications or overlapping datasets were also excluded to prevent redundancy in the synthesis of findings.

### Searching Strategy

A comprehensive search strategy was employed to identify relevant studies on Diabetes Self-Management Education and Support (DSMES). The primary sources of data were six full-text articles provided in the uploaded files, which were carefully screened and included as the core evidence base of this review. To complement these, additional literature searches were conducted across major electronic databases including Scopus, PubMed, Web of Science (WoS), Science Direct, and Ebsco. The search was designed to capture studies published between 2018 and 2025, with a particular focus on the years 2020 to 2025 in order to ensure that the discussion was informed by the most recent developments in the field.

The search terms combined keywords and Medical Subject Headings (MeSH) related to “diabetes self-management education,” “DSMES,” “type 2 diabetes,” “self-care,” “telemedicine,” “culturally tailored,” and “learning model.” Boolean operators (AND, OR) were used to broaden or narrow the scope of the search, depending on the combination of terms. Titles and abstracts retrieved from the databases were screened to identify potential studies, and the full texts of relevant articles were then assessed against the predefined inclusion and exclusion criteria.

To ensure comprehensiveness, the reference lists of included studies and related reviews were also screened manually to identify additional eligible publications. This snowballing technique helped to minimize the risk of missing relevant literature. Duplicates across databases were removed before screening to avoid redundancy. The overall search process and screening followed the PRISMA 2020 guidelines, with a structured flow of identification, screening, eligibility assessment, and inclusion documented in the PRISMA flow diagram.

### Procedure of Data Extraction

The procedure of data extraction was carried out systematically using a standardized form developed for this review. For each included study, key information was independently extracted by the reviewers to ensure accuracy and consistency. The data extraction process captured bibliographic details (title, author, year of publication, and country of study), study characteristics (design, sample size, population characteristics such as age and gender), and details of the intervention (type of DSMES, duration, frequency, and delivery mode, e.g., face-to-face, group-based, culturally adapted, or telemedicine-supported). Where applicable, comparator conditions such as usual care or alternative interventions were also recorded.

Outcome measures were carefully extracted to reflect the primary aims of DSMES interventions. These included clinical outcomes such as HbA1c, incidence of diabetic foot complications, and other metabolic indicators; psychosocial outcomes such as diabetes distress, well-being, and quality of life; and behavioral outcomes such as self-care practices and adherence to treatment regimens. For studies reporting quantitative data, numerical results including means, standard deviations, confidence intervals, and p-values were documented. For qualitative or developmental studies, descriptions of model development, feasibility, and patient acceptability were summarized.

To ensure data integrity, extracted information was cross-checked between reviewers. Any discrepancies in interpretation were discussed and resolved by consensus. When necessary, supplementary information was obtained from appendices, supplementary materials, or references cited within the articles. The extracted data were subsequently tabulated to provide a structured comparison across studies, which informed the synthesis presented in the Results section. This systematic approach ensured that the findings were robust, transparent, and aligned with the methodological standards recommended by PRISMA 2020.

## RESULT

The results of the research articles on the impact of culture-based family empowerment on anemia prevention behavior in pregnant women are presented in Table 1.

**Table 1.**
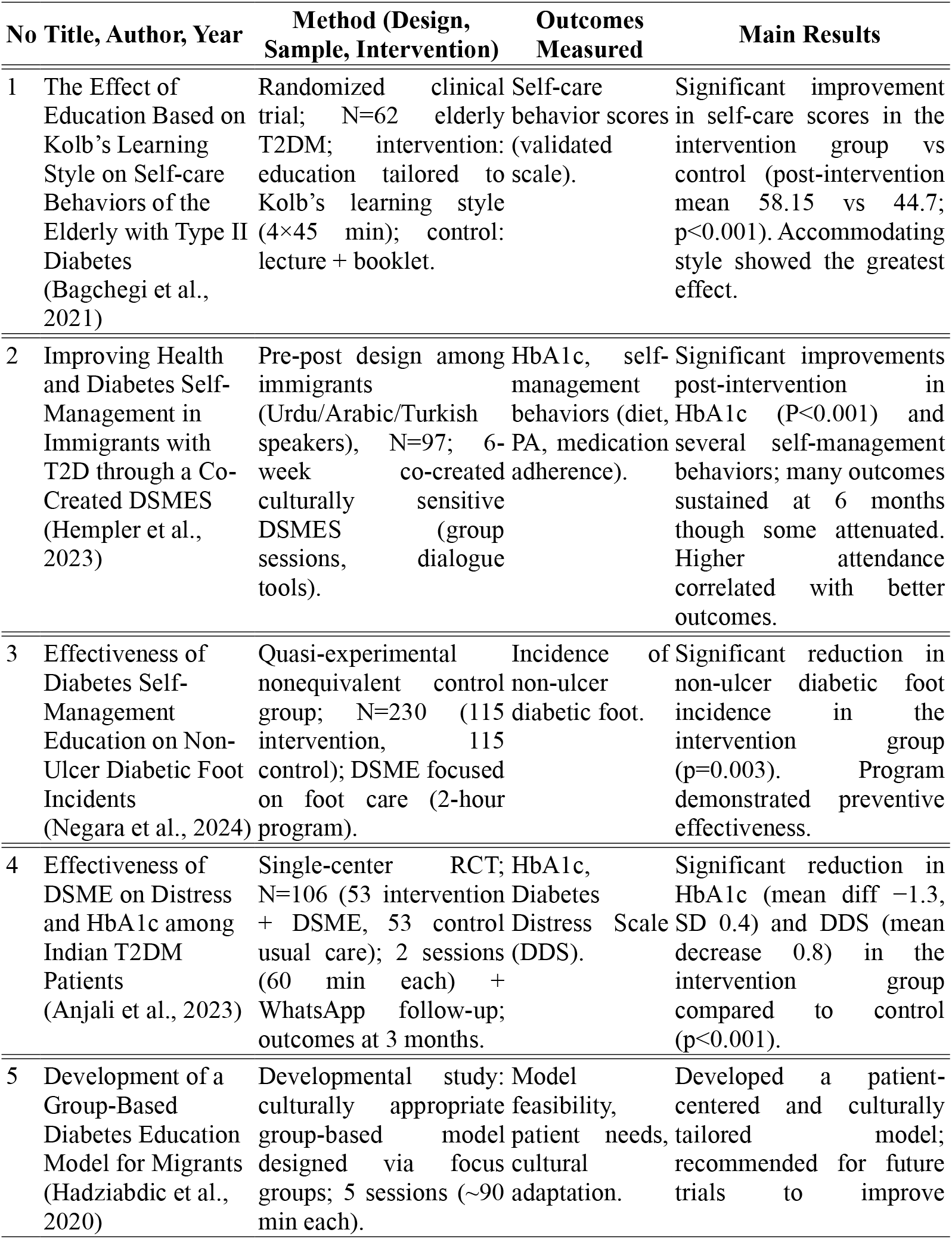
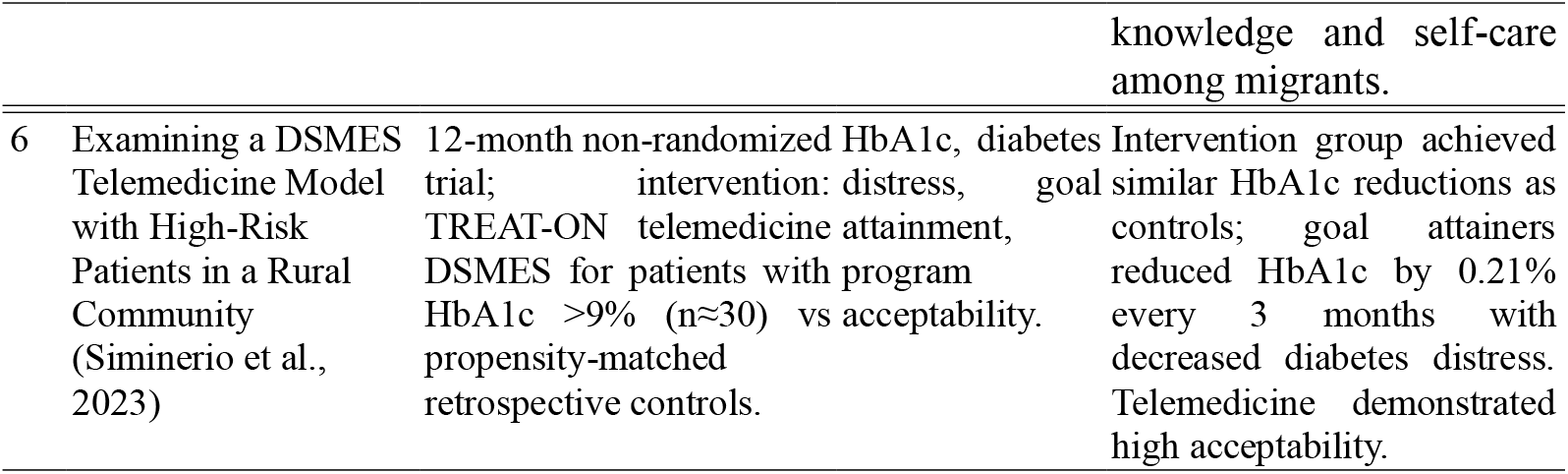
The research articles on Diabetes Self-Management Education and Support Models and Their Impact on Clinical and Psychosocial Outcomes

## Discussion

The findings from the six studies analyzed in this review consistently affirm the effectiveness of Diabetes Self-Management Education and Support (DSMES) in improving various aspects of self-care among patients with type 2 diabetes. Although variations existed in study design, duration, and intervention contexts, all studies reported significant improvements in at least one primary outcome.

Personalized education through Kolb’s learning style approach demonstrated a clear impact on self-care behaviors. Bagchegi *et al* (2021) reported that self-care scores in the intervention group increased from a mean of 46.1 (pre-intervention) to 58.15 post-intervention, whereas the control group improved only slightly from 43.9 to 44.7. The between-group difference was statistically significant (p < 0.001), with the accommodating learning style showing the strongest response. These results align with the health education literature emphasizing that experiential learning methods tailored to individual learning styles enhance knowledge retention and behavioral application (Davitadze *et al*., 2022).

Culturally tailored interventions also demonstrated strong positive effects. In a six-week co-created program for immigrant communities, Hempler *et al* (2023) found a mean reduction in HbA1c of 0.41% (p < 0.001) at three months, along with improvements in self-management behaviors such as dietary adherence and physical activity. Although some gains diminished by six months, self-care scores remained higher than baseline. Similarly, Hadziabdic *et al*., (2020) highlighted the potential of culturally adapted group education models, although their study focused on model development without clinical outcomes. These findings are consistent with Zhang (2024), who reported that culturally sensitive DSMES programs are more effective among migrant and ethnic minority populations compared with standard approaches.

Telemedicine-based interventions are increasingly relevant, particularly for high-risk populations and those in rural areas. Siminerio et al. (2023), through the TREAT-ON model, demonstrated that patients with baseline HbA1c >9% who received telehealth DSMES achieved mean HbA1c reductions of 0.21% every three months. Furthermore, diabetes distress scores decreased significantly, and patients reported high satisfaction with the flexibility of the service. These findings align with the meta-analysis by Usefi et al. (2024), which showed that online DSMES interventions reduced HbA1c by an average of −0.37% (95% CI: −0.52 to −0.21) over 3–6 months, as well as with Chiaranai et al. (2024), who highlighted the effectiveness of telehealth in maintaining glycemic control during the COVID-19 pandemic. Notably, the study from India by Anjali et al. (2023) demonstrated substantial clinical benefits. Patients who received two DSMES sessions (60 minutes each) with follow-up via WhatsApp experienced a mean HbA1c reduction of 1.3% (SD 0.4; p < 0.001), while the control group showed no meaningful change. Moreover, Diabetes Distress Scale (DDS) scores decreased by a mean of 0.8 points in the intervention group, reflecting reduced emotional burden related to diabetes management. This reduction is clinically relevant, as persistent distress can undermine adherence and worsen long-term outcomes.

Beyond glycemic and psychosocial outcomes, DSMES also proved effective in preventing complications. Negara et al. (2024) reported that a DSMES program focused on foot care significantly reduced the incidence of non-ulcer diabetic foot (p = 0.003) in a quasi-experimental study involving 230 patients. The intervention group had markedly lower complication rates compared to controls. This finding supports evidence from Ju *et al* (2023), who showed that nurse-led foot care education can reduce the risk of diabetic ulcers by enhancing patients’ practical skills.

Despite these benefits, challenges remain regarding the sustainability of intervention effects. Hempler et al. (2022) noted that several behavioral improvements diminished at six-month follow-up, though they remained above baseline levels. This underscores the need for ongoing support, whether through telemedicine, peer groups, or structured follow-up in primary care. Similarly, Hjelm and Hadziabdic (2025) observed that DSMES effects tend to be strongest within the first 3-6 months but may wane without consistent reinforcement.

Overall, evidence from the six studies analyzed supplemented by recent literature indicates that structured, patient centered DSMES interventions, particularly those personalized through learning styles, culturally adapted, and supported by technology, yield significant improvements in glycemic control, distress, self-care, and complication prevention. However, study heterogeneity and the lack of long-term data remain key limitations. Further research through large-sample randomized controlled trials with durations of ≥12 months, alongside cost-effectiveness analyses, is needed to confirm sustainability and scalability of DSMES programs across diverse healthcare settings.

## CONCLUSION

This systematic review demonstrates that structured Diabetes Self-Management Education and Support (DSMES) interventions are effective in improving glycemic control, self-care behaviors, psychological outcomes, and prevention of complications in adults with type 2 diabetes. Personalized approaches based on learning styles, culturally tailored programs, and telemedicine-supported models showed the most consistent benefits. However, long-term sustainability of these effects remains a challenge, underscoring the need for ongoing support and larger randomized controlled trials to evaluate cost-effectiveness and durability of different DSMES delivery models.

## Data Availability

All data produced in the present work are contained in the manuscript

## Notes

### Competing Interest Statement

The authors have declared no competing interest.

### Funding Statement

This study did not receive any funding

